# Cardiac Myosin Inhibitors in Adults with Symptomatic Non-Obstructive Hypertrophic Cardiomyopathy: A Systematic Review and Meta-analysis of Randomized Controlled Trials

**DOI:** 10.64898/2026.09.08.26362195

**Authors:** Jan Koelemen, David Koeckerling, Steffen Rosskopf, Christoph Reich, Ali Amr, Elham Kayvanpour, Norbert Frey, Farbod Sedaghat-Hamedani, Benjamin Meder

## Abstract

**Background:** Cardiac myosin inhibitors (CMIs) directly target sarcomeric hypercontractility and have demonstrated clinical efficacy in obstructive hypertrophic cardiomyopathy (HCM). In symptomatic non-obstructive HCM (nHCM), the phase 3 randomized controlled trials (RCT) differed in whether their primary efficacy endpoints reached statistical significance, although the direction and magnitude of treatment effects were broadly comparable. We therefore conducted a systematic review and meta-analysis to synthesize the randomized evidence on the efficacy and safety of CMIs in symptomatic nHCM.

**Methods:** We systematically searched for randomized placebo-controlled trials evaluating CMIs in adults with symptomatic nHCM according to PRISMA guidance and a prospectively registered protocol (PROSPERO CRD420261466576). Primary efficacy outcomes were changes from baseline in peak oxygen uptake (pVO_2_) and Kansas City Cardiomyopathy Questionnaire Clinical Summary Score (KCCQ-CSS), assessed at each trial’s primary endpoint. Secondary outcomes included improvement by at least 1 New York Heart Association (NYHA) functional class and change in N-terminal pro-B-type natriuretic peptide (NT-proBNP). The safety outcome was LVEF <50%. Random-effects models were used to pool mean differences (MDs), risk ratios (RRs), and ratios of geometric mean changes.

**Results:** Three RCTs comprising 1,156 participants were included: MAVERICK-HCM, ODYSSEY-HCM, and ACACIA-HCM. Compared with placebo, CMIs were associated with improvements in pVO_2_ (MD, 0.55 mL/kg/min; 95% CI, 0.22-0.87; I^2^=0%; P=0.001) and KCCQCSS (MD, 2.71 points; 95% CI, 0.87-4.55; I^2^=0%; P=0.004). Improvement by at least 1 NYHA functional class was more frequent with CMIs (RR, 1.29; 95% CI, 1.03-1.62; I^2^=24.3%; P=0.024). Across the two phase 3 trials, CMIs reduced NT-proBNP (ratio of geometric mean changes, 0.42; 95% CI, 0.39-0.46; I^2^=0%; P<0.001), while LVEF <50% occurred more frequently with CMIs (RR, 12.77; 95% CI, 5.98-27.27; I^2^=0%; P<0.001).

**Conclusions:** In this meta-analysis of randomized trials in symptomatic nHCM, CMIs’ pooled estimates favoured CMIs over placebo for improvements in exercise capacity, health status, and NYHA functional class, together with a marked reduction in NT-proBNP. These potential benefits were accompanied by an increased risk of LVEF reduction below 50%.

## Introduction

Hypertrophic cardiomyopathy (HCM) is characterized by sarcomeric hypercontractility and impaired myocardial relaxation. Cardiac myosin inhibitors (CMIs), which directly target sarcomeric hypercontractility, have transformed the treatment of obstructive HCM (oHCM). However, no targeted pharmacological therapy is currently approved for symptomatic nonobstructive HCM (nHCM) (1-8).

The shared underlying sarcomeric pathophysiology provides a rationale for extending myosin inhibition to nHCM (3,9). In the phase 3 ODYSSEY-HCM trial, at week 48 mavacamten markedly reduced N-terminal pro-B-type natriuretic peptide (NT-proBNP) levels, but improvements in peak oxygen uptake (pVO_2_) (+0.47 mL/kg/min; 95% CI: -0.03 to 0.98; P=0.07) and Kansas City Cardiomyopathy Questionnaire Clinical Summary Score (KCCQ-CSS) (+2.7 points; 95% CI: -0.1 to 5.6; P=0.06) narrowly missed statistical significance, while reductions in left ventricular ejection fraction (LVEF) <50% occurred statistically more frequent with mavacamten than placebo (21.5% vs 1.7%) (10). The recently published phase 3 ACACIAHCM trial demonstrated at week 36 in addition to comparable biomarker response significant improvements with aficamten in both pVO_2_ (+0.67 mL/kg/min; 95% CI: 0.22-1.11; P=0.003) and KCCQ-CSS (+3.0 points; 95% CI: 0.5-5.5; P=0.02), with LVEF <50% occurring in 10.5% versus 0.8% with placebo (11). Despite the differences in reaching statistical significance, the numerical effects between both studies having very similar study design, inclusion and exclusion criteria, seem quite comparable. To evaluate the efficacy and safety of CMIs as a therapeutic class in symptomatic nHCM we conducted a systematic review and meta-analysis.

## Methods

This systematic review and meta-analysis is reported according to the Preferred Reporting Items for Systematic Reviews and Meta-Analyses (PRISMA) guidance, with preregistration on PROSPERO (CRD420261466576) (12,13). The search syntax was developed with the help of a medical librarian (V.B.). CENTRAL was systematically searched from inception through August 29, 2026 for randomized placebo-controlled trials evaluating cardiac myosin inhibitors in adults with symptomatic nonobstructive hypertrophic cardiomyopathy (Supplementary Material). Additional pertinent studies were sought through backward citation searching and screening of clinical trial registries and proceedings from major cardiology conferences. Two reviewers independently performed abstract screening, full-text review and data extraction, with disagreements resolved by consensus. Risk of bias was assessed using the Cochrane Risk of Bias 2 (ROB 2) tool for randomized trials (14).

The prespecified primary efficacy outcomes were changes from baseline in pVO_2_ and KCCQ-CSS, evaluated at the respective primary endpoint time point of each trial. Secondary outcomes included improvement by at least 1 New York Heart Association (NYHA) functional class and change in NT-proBNP. The prespecified safety outcome was the proportion of participants experiencing at least one postbaseline LVEF measurement <50%. The principal summary metrics were mean differences on the original scale for continuous outcomes and risk ratios (RRs) for dichotomous outcomes, with associated 95% confidence intervals. Adjusted estimates corresponding most closely to the intention-to-treat principle were preferred for efficacy endpoints, the safety endpoint was based on the trial-defined safety populations. NT-proBNP ratios of geometric mean changes were analyzed on the logarithmic scale and exponentiated for presentation. Summary metrics were pooled using inverse-variance weighting, with random-effects models fitted using restricted maximum likelihood estimation. Common-effect models were fitted as sensitivity analysis. Two-sided P values <0.05 were considered statistically significant. Heterogeneity was assessed using Cochran Q, I^2^, and τ^2^. Analyses were performed using R (Version 4.5.2) and the metafor package. Formal assessment of small-study effects and meta-regression were not performed because of the small number of eligible trials.

## Results

Only randomized placebo-controlled trials on CMIs in nHCM were included: the phase 2 MAVERICK-HCM trial and the phase 3 ODYSSEY-HCM and ACACIA-HCM trials, comprising 1,156 participants overall. MAVERICK-HCM (n=59) and ODYSSEY-HCM (n=580) evaluated mavacamten, whereas ACACIA-HCM (n=517) evaluated aficamten. Main baseline characteristics of the study cohorts are described in Supplementary Table 1. Risk of bias was low across all included trials according to the Cochrane RoB 2 tool.

Cardiac myosin inhibition significantly improved pVO_2_ compared with placebo (mean difference (MD): 0.55 mL/kg/min; 95% CI: 0.22-0.87; I^2^=0%; P=0.001) and KCCQ-CSS (MD: 2.71 points; 95% CI: 0.87-4.55; I^2^=0%; P=0.004) (Figure 1 A & B). Improvement by at least 1 NYHA functional class was also more frequent with cardiac myosin inhibition (RR: 1.29; 95% CI: 1.03-1.62; I^2^=24.3%; P=0.024) (Figure 1 C; Supplemental Figure 1). Results were consistent in common-effect models (Figure 1 A & B; Supplementary Figures 1-3).

**Figure 1.**
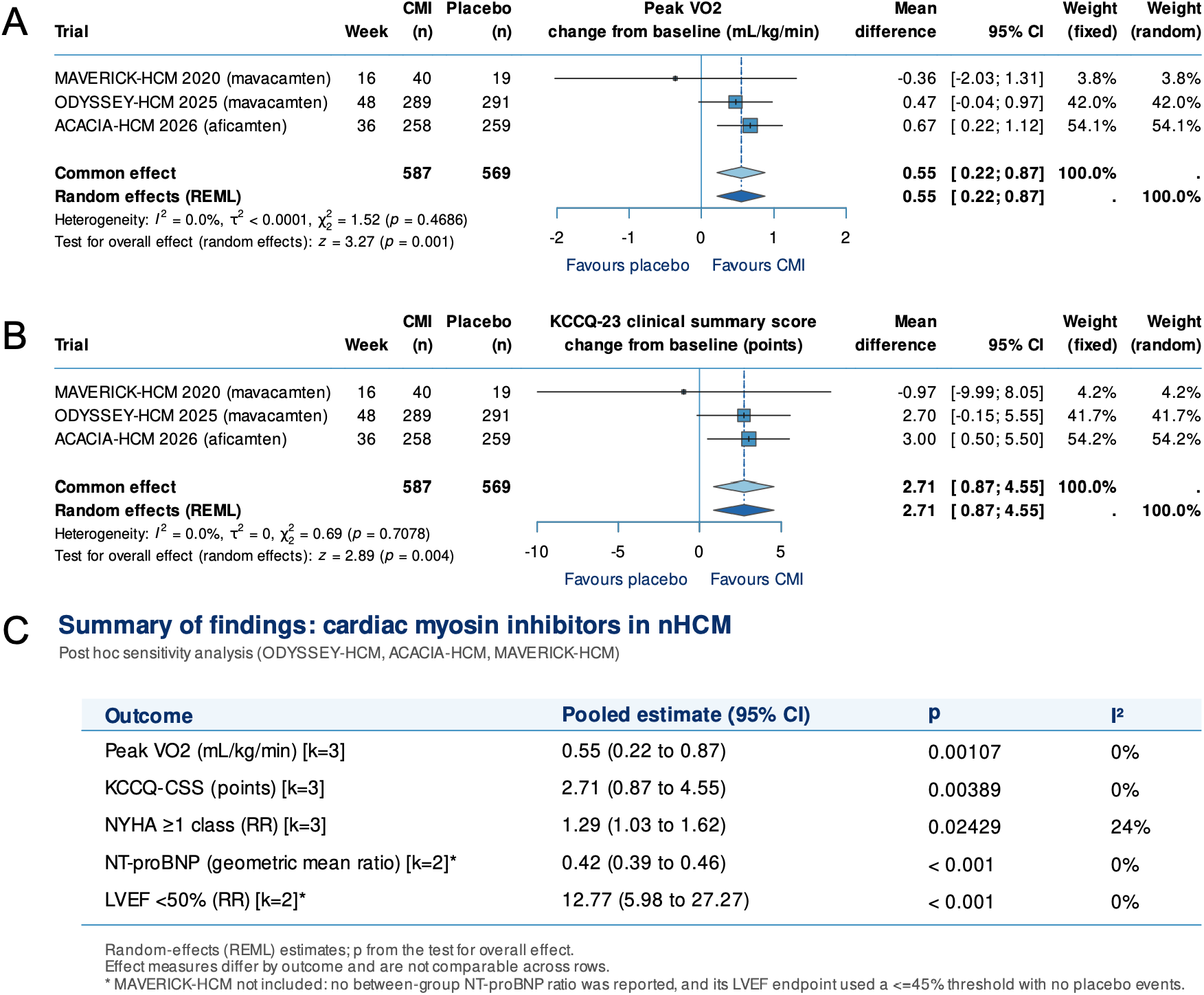
Summary of findings: meta-analysis of cardiac myosin inhibitors in nonobstructive hypertrophic cardiomyopathy. Forest plots show pooled effects of cardiac myosin inhibitors (CMIs) versus placebo on **(A)** peak oxygen uptake (peak VO_2_) and **(B)** Kansas City Cardiomyopathy Questionnaire Clinical Summary Score (KCCQCSS). **(C)** Summary of pooled efficacy, biomarker, and safety outcomes. Random-effects models used restricted maximum likelihood (REML), common-effect estimates are shown for comparison. NT-proBNP and LVEF <50% analyses included ODYSSEY-HCM and ACACIA-HCM only. CI indicates confidence interval; LVEF, left ventricular ejection fraction; NT-proBNP, N-terminal pro-B-type natriuretic peptide; NYHA, New York Heart Association.

For NT-proBNP and LVEF <50%, MAVERICK-HCM was excluded because comparable effect estimates were not available. Cardiac myosin inhibition reduced NT-proBNP, with a pooled ratio of geometric mean changes of 0.42 (95% CI: 0.39-0.46; I^2^=0%; P<0.001), corresponding to an approximately 58% greater reduction relative to placebo (Figure 1 C; Supplementary Figure 2). Regarding safety, LVEF decline <50% occurred more frequently with cardiac myosin inhibition (RR: 12.77; 95% CI: 5.98-27.27; I^2^=0%; P<0.001) (Figure 1 C; Supplemental Figure 3).

## Discussion

In this meta-analysis of 3 randomized trials, pooled estimates favoured cardiac myosin inhibitors over placebo with regards to improvements of exercise capacity and health status in patients with symptomatic nonobstructive HCM. CMIs also resulted in a marked reduction in NT-proBNP, supporting a strong biological effect on myocardial wall stress.

The rationale for considering CMIs at the class level is supported by their shared mechanism and the concordant biological responses observed across trials. Both agents reduce excessive actin-myosin interaction and thereby target sarcomeric hypercontractility, a fundamental disease mechanism irrespective of left ventricular outflow tract obstruction. In the official German AMNOG early benefit assessment procedure, statistical comparisons with the methodology of Bucher (15) between mavacamten and aficamten pointed to similar efficacy metrics of both CMIs in oHCM (16). The pooled results in our meta-analysis on non-obstructive HCM now place the apparently discordant primary results of ODYSSEY-HCM and ACACIAHCM into a broader context. ODYSSEY-HCM did not meet the co-primary efficacy endpoints, although treatment effects numerically favored mavacamten for both pVO_2_ (between-group difference: 0.47 mL/kg/min; 95% CI: -0.03 to 0.98; P=0.07) and KCCQ-CSS (2.7 points; 95% CI: -0.1 to 5.6; P=0.06) (10). Importantly, biological target engagement was evident, with NT-proBNP decreasing by 59% at 48 weeks (10). ACACIA-HCM subsequently demonstrated improvements in both pVO_2_ (0.67 mL/kg/min; 95% CI: 0.22-1.11; P=0.003) and KCCQ-CSS (3.0 points; 95% CI: 0.5-5.5; P=0.02), accompanied by a similarly pronounced reduction in NT-proBNP (11). Thus, the two phase 3 trials did not demonstrate fundamentally divergent biological or clinical responses. Rather, effect estimates for both primary efficacy outcomes were directionally concordant, while only ACACIA-HCM crossed conventional thresholds of statistical significance.

The pooled risk of LVEF <50% was increased with cardiac myosin inhibition (RR: 12.77; 95% CI: 5.98-27.27; I^2^=0%; P<0.001). Importantly, reductions in LVEF were managed differently between trials. In ODYSSEY-HCM, LVEF <50% mandated treatment interruption, and 21,5% of mavacamten-treated patients required interruption because of systolic dysfunction vs. 1,7% with placebo (10). In ACACIA-HCM, LVEF <50% occurred in 10.5% receiving Aficamten vs. 0,8% with placebo, but reductions could generally be managed by dose reduction (11). Treatment interruption was mandated for LVEF <40% and occurred in 2.7% (11). These differences in LVEF-guided dose adjustment may have influenced cumulative treatment exposure and thereby observed treatment effects. However, this remains hypothesis-generating because cross-trial comparisons cannot establish whether these differences explain the observed efficacy results.

## Limitations

This analysis has several limitations. Only 3 randomized trials were available, limiting the precision of heterogeneity estimates and precluding meaningful assessment of treatment-effect modifiers or differences between individual agents. However, the designs of the two phase 3 trials were remarkably comparable and also the baseline characteristics suggest relatively good homogeneity. NT-proBNP and LVEF <50% could be pooled only from the 2 phase 3 trials because MAVERICK-HCM did not provide a directly comparable NT-proBNP effect estimate and reported systolic dysfunction using an LVEF threshold of ≤45%. The timing of primary endpoint assessment varied across trials (16, 36, and 48 weeks), introducing potential temporal heterogeneity; however, data from oHCM trials and registries suggest that the largest treatment effects emerge within the first 3 months (6,7,17,18). The trials also differed in pharmacokinetic properties and dose-adjustment strategies, and aggregate trial-level data precluded assessment of whether treatment exposure or LVEF-guided dose modification mediated efficacy.

## Conclusion

Pooled randomized evidence indicates that cardiac myosin inhibition improves exercise capacity and health status and reduces myocardial wall stress in symptomatic nHCM. The consistent direction of functional effects and concordant biological responses across trials support targeting sarcomeric hypercontractility as a therapeutic strategy beyond obstructive HCM. However, the magnitude of clinical benefit and the increased risk of systolic dysfunction underscore the need to optimize treatment exposure and identify patient subgroups at greatest risk of LVEF decline.

## Data Availability

All data underlying this study were derived from publicly available published articles and their supplementary materials. No individual participant-level data were used or generated.

## Abbreviations

CENTRAL: Cochrane Central Register of Controlled Trials
CI: confidence interval
CMI: cardiac myosin inhibitor
GRADE: Grading of Recommendations Assessment, Development and Evaluation
HCM: hypertrophic cardiomyopathy
KCCQ-CSS: Kansas City Cardiomyopathy Questionnaire Clinical Summary Score
LVEF: left ventricular ejection fraction
MD: mean difference
nHCM: nonobstructive hypertrophic cardiomyopathy
NT-proBNP: N-terminal pro-B-type natriuretic peptide
NYHA: New York Heart Association
oHCM: obstructive hypertrophic cardiomyopathy
PRISMA: Preferred Reporting Items for Systematic Reviews and Meta-Analyses
PROSPERO: International Prospective Register of Systematic Reviews
pVO_2_: peak oxygen uptake
RR: risk ratio

## Acknowledgements

CR is funded by the Clinician Scientist Program of Heidelberg University, Faculty of Medicine. We thank Volker Braun (medical librarian) for his assistance in developing and conducting the systematic literature search strategy. All content was reviewed, validated, and approved by the authors, who take full responsibility for the accuracy and integrity of the work.

