## Supplementary Material for "Cardiac Myosin Inhibitors in Adults with Symptomatic Non-Obstructive Hypertrophic Cardiomyopathy: A Systematic Review and Meta-analysis of Randomized Controlled Trials"

#### Supplementary methods

##### Detailed search strategy

The electronic search strategy was developed around 2 concepts: cardiac myosin inhibition and nonobstructive hypertrophic cardiomyopathy. The cardiac myosin inhibitor concept included the generic terms myosin inhibitor and cardiac myosin inhibitor, as well as the individual agents mavacamten and aficamten. The disease concept included hypertrophic cardiomyopathy in combination with the terms nonobstructive and non-obstructive. Searches were performed in the Cochrane Central Register of Controlled Trials (CENTRAL) using title (ti), abstract (ab), and keyword (kw) fields. Proximity operators were used to account for variations in terminology, and truncation was applied to capture alternative word endings. The 2 concepts were combined using the Boolean operator AND. The complete CENTRAL search syntax was: ((myosin NEAR/2 inhibitor*) OR Mavacamten OR aficamten):ti,ab,kw AND ((hypertrophic NEAR/2 (nonobstructive OR non-obstructive) NEXT cardiomyopath*):ti,ab,kw). No language restrictions were applied.

#### Supplementary Tables

**Supplementary Table 1. Baseline Characteristics of Included Trials**

| Variable | MAVERICK-HCM | ODYSSEY-HCM | ACACIA-HCM |
|---|---|---|---|
| Eligibility criteria | Adults ≥18 years with symptomatic nHCM (NYHA II-III); NT-proBNP >300 pg/mL; LVEF ≥55%; LV wall thickness ≥15 mm or ≥13 mm with family history of HCM; resting or provocable LVOT/intracavitary gradient ≤30 mm Hg. | Adults ≥18 years with symptomatic nHCM (NYHA II-III); KCCQ-CSS ≤85; LVEF ≥60%; LVOT gradient <30 mm Hg at rest and <50 mm Hg with provocation; RER ≥1.0 and elevated NT-proBNP. | Adults with symptomatic nHCM (NYHA II-III); KCCQ-CSS ≤85; LVEF ≥60%; NT-proBNP ≥300 pg/mL; LV wall thickness ≥15 mm or ≥13 mm with a disease-causing variant or family history of HCM; resting LVOT gradient <30 mm Hg and Valsalva gradient <50 mm Hg. |
| Study size, n | 59 | 580 | 517 |
| CMI group, n | 40 | 289 | 258 |
| Placebo group, n | 19 | 291 | 259 |
| Age, mean ± SD, years | 53.9 ± 15.7 | 56.0 ± 14.5 | 55.1 ± 16.1 |
| Female sex, n (%) | 34 (57.6%) | 266 (45.9%) | 277 (53.6%) |
| Peak $VO_2$, mean ± SD, mL/kg/min | 19.6 ± 5.8 | 18.1 ± 5.7 | 18.0 ± 5.2 |
| KCCQ-CSS, mean ± SD, points | n.a. | 56.9 ± 19.9 | 65.6 ± 15.8 |
| NT-proBNP, ng/L | CMI 821 (790-1293); placebo 914 (770-1558) (geometric mean (95% CI)) | 918 (463-1725) (median, IQR, ng/L) | 893 (n.a.) (median, ng/L) |
| hs-cTnI, median, ng/L | n.a. | CMI 29.5 (14.1-81.1); placebo 28.1 (14.4-110.9) | CMI 30 (15-77); placebo 28 (14-113) |
| LVEF, mean ± SD, % | 68.0 ± 6.3 | 65.7 ± 4.0 | 68.2 ± 3.7 |
| LVOT gradient, mean ± SD, mm Hg | CMI 8.8 ± 3.5; placebo 7.8 ± 2.5 (peak gradient) | 10.7 ± 6.8 (Valsalva) | 11.6 ± 8.6 (Valsalva) |
| Maximum LV wall thickness, mean ± SD, mm | 20.0 ± 3.9 | 20.8 ± 4.1 | 22.7 ± 5.5 |
| NYHA class I, n (%) | 0 (0.0%) | 0 (0.0%) | 5 (1.0%) |
| NYHA class II, n (%) | 46 (78.0%) | 405 (69.8%) | 354 (68.5%) |
| NYHA class III, n (%) | 13 (22.0%) | 175 (30.2%) | 158 (30.6%) |
| NYHA class IV, n (%) | 0 (0.0%) | 0 (0.0%) | 0 (0.0%) |
| Arterial hypertension, n (%) | n.a. | 260 (44.8%) | 202 (39.1%) |
| Diabetes mellitus, n (%) | n.a. | 69 (11.9%) | 63 (12.2%) |
| Prior septal reduction therapy, n (%) | n.a. | 34 (5.9%) | 60 (11.6%) |
| Family history of HCM, n (%) | n.a. | 251 (43.3%) | 211 (40.8%) |
| History of atrial fibrillation, n (%) | n.a. | 180 (31.0%) | 185 (35.8%) |
| Beta-blocker use, n (%) | 37 (62.7%) | 453 (78.1%) | 400 (77.4%) |
| Calcium-channel blocker use, n (%) | 13 (22.0%) | 70 (12.1%) | 70 (13.5%) |
| Disopyramide use, n (%) | n.a. | 17 (2.9%) | n.a. |

Values are mean ± SD, median (IQR), geometric mean (95% CI), or n (%), as indicated. NT-proBNP in MAVERICK-HCM is reported separately by randomized group as geometric mean (95% CI). LVOT gradient definitions differed among trials. CMI = cardiac myosin inhibitor; HCM = hypertrophic cardiomyopathy; hs-cTnI = high-sensitivity cardiac troponin I; KCCQ-CSS = Kansas City Cardiomyopathy Questionnaire Clinical Summary Score; LVEF = left ventricular ejection fraction; LV = left ventricular; LVOT = left ventricular outflow tract; NT-proBNP = N-terminal pro-B-type natriuretic peptide; NYHA = New York Heart Association; RER = respiratory exchange ratio; SD = standard deviation; IQR = interquartile range; n.a. = not available/not reported.

### Supplementary Figures

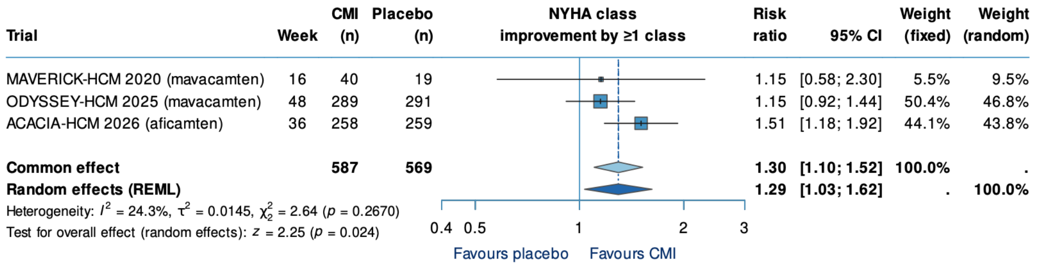


***Supplementary Figure 1.*** **Effect of cardiac myosin inhibitors on NYHA functional class in nonobstructive hypertrophic cardiomyopathy.** Forest plot showing the pooled risk ratio for improvement by at least 1 NYHA functional class with cardiac myosin inhibitors (CMIs) versus placebo. Random-effects estimates were calculated using restricted maximum likelihood (REML), common-effect estimates are shown for comparison. CI indicates confidence interval; NYHA, New York Heart Association.

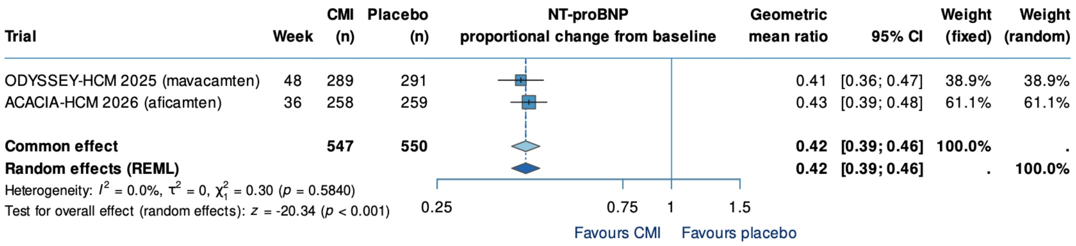


***Supplementary Figure 2.*** **Effect of cardiac myosin inhibitors on NT-proBNP in nonobstructive hypertrophic cardiomyopathy**. Forest plot showing the pooled geometric mean ratio for proportional change from baseline in N-terminal pro-B-type natriuretic peptide (NT-proBNP) with CMIs versus placebo. Random-effects estimates were calculated using REML, common-effect estimates are shown for comparison. CI indicates confidence interval.

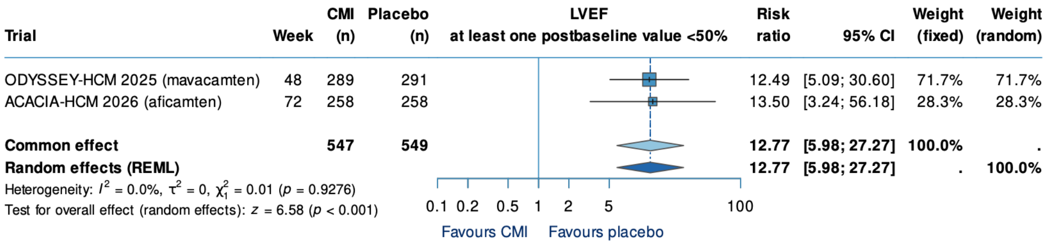


***Supplementary Figure 3.*** **Effect of cardiac myosin inhibitors on left ventricular systolic function in nonobstructive hypertrophic cardiomyopathy.** Forest plot showing the pooled risk ratio for at least 1 postbaseline left ventricular ejection fraction (LVEF) <50% with CMIs versus placebo. Random-effects estimates were calculated using REML, common-effect estimates are shown for comparison. CI indicates confidence interval; LVEF, left ventricular ejection fraction.
